# Site-specific DNA methylation in KCNJ11 promoter contributes to type 2 diabetes

**DOI:** 10.1101/2024.07.13.24310360

**Authors:** Mengmeng Zhu, Qiaoliang Huang, Heng Li, Yujie Zhao, Heming Guo, Tao Wang, Xiaodan Liu, Yun Huang, Ji Hu, Chen Fang, Jian Huang

**Author notes:** These authors contributed equally to this work (Mengmeng Zhu, Qiaoliang Huang). Corresponding authors: **Fang, C. E-mail:**, **Huang, J. E-mail:**.

## Abstract

**Aims:** This study explores the correlation between site-specific methylation levels of the KCNJ11 promoter and type 2 diabetes mellitus (T2DM), analyzing potential molecular mechanisms.

**Methods:** Thirty patients newly diagnosed with T2DM and 30 healthy controls were selected to determine the CpG methylation levels in the promoter region of the KCNJ11 gene using the bisulfite assay. The online software JASPAR was used to predict transcription factors binding to differentially methylated sites. Key transcription factors were further validated through quantitative PCR (q-PCR) and chromatin immunoprecipitation followed by PCR (ChIP-PCR).

**Results:** Methylation at multiple CpG sites within the KCNJ11 gene promoter was significantly reduced in newly diagnosed T2DM patients compared to healthy individuals. The methylation status of CpG-471, a site crucial for the binding of the transcription factor TCF12, emerged as potentially influential in T2DM pathogenesis. This reduction in methylation at CpG-471 may enhance TCF12 binding, thereby altering KCNJ11 expression.

**Conclusion:** Hypomethylation of specific CpG sites in the promoter region of the KCNJ11 gene in patients with incipient T2DM potentially contributes to the disease’s pathogenesis. This hypomethylation enhances the transcriptional activity of TCF12, which binds to these sites, possibly influencing regulatory pathways involved in pancreatic beta-cell function.

## 1. Introduction

Type 2 diabetes mellitus (T2DM) is a complex polygenic disease[1], primarily driven by insulin resistance and inadequate insulin production from β cells[2]. According to the updated version of the International Diabetes Federation (IDF) Diabetes Atlas, more than 500 million patients worldwide have diabetes, and this number is expected to increase by 20% by 2030 and 46% by 2045[3]. China has the highest prevalence of diabetes and prediabetes, with an estimated 10.9% of Chinese adults diagnosed with diabetes and 35.7% with prediabetes[4]. Due to the significant increase in the incidence of T2DM and its concomitant acute and chronic complications, it is an important cause of patient death and increased healthcare costs, which brings a huge economic burden to the society, and there is an urgent need to actively intervene in the treatment of T2DM from the etiologic and early prevention perspectives[5, 6]. The etiology of T2DM is complex, resulting from a combination of genetic and environmental factors[7]. An important area of current research is the search for reliable, sensitive and easily accessible biomarkers for T2DM. In this context, a growing number of studies[8–13]suggest that genetic and epigenetic mechanisms together play an important role in the pathogenesis of T2DM patients.

Differential variability in DNA methylation, often indicative of epigenetic dysregulation, is associated with various phenotypes, pathologies, and adverse environments[9]. Initial epigenetic studies in pancreatic islets and skeletal muscle of T2DM patients identified altered methylation patterns[14].Recent genome-wide DNA methylation studies have confirmed these findings, revealing altered methylation levels in diabetes-associated genes like TCF7L2[15], PDX-1[16], CDKN1A[17], FTO[18], and PPARGC1A[19], with changes in their promoter regions correlating with diabetes risk and severity. Additionally, the KCNJ11 gene, encoding the Kir6.2 subunit of the ATP-sensitive K+ channel crucial for insulin secretion, has been linked to T2DM development through both genetic mutations and methylation changes[20–28]. Research has shown that methylation patterns of KCNJ11 vary by geography and ethnicity, with significant differences observed between Indian and European T2DM populations[29]. These findings highlight the complex interplay of genetic and epigenetic factors in T2DM, emphasizing the potential of methylation studies in understanding and addressing this widespread disease.

While genome-wide association studies have extensively documented changes in methylation patterns, few have focused on methylation at specific sites. Therefore, our study aims to investigate the correlation between specific methylation sites in the KCNJ11 gene promoter and the onset of Type 2 Diabetes Mellitus (T2DM), as well as how these modifications affect gene expression through transcription factor regulation. Supporting literature suggests that age-related methylation changes detectable in blood could serve as biomarkers for T2DM[30–32]. To this end, we selected 30 patients newly diagnosed with T2DM (20 men and 10 women), average age 57.0 ± 13.1 years. We also recruited 30 matched controls—18 males and 12 females, average age 56.7 ± 10.1 years, all meeting specific criteria for glucose tolerance. Exclusion criteria were a prior diabetes diagnosis, serious concurrent conditions, pregnancy, and dropout from follow-up. Comparisons between baseline data of the two groups showed no significant differences (P > 0.05), confirming their comparability. Preliminary results indicate significantly decreased methylation at several sites within the KCNJ11 promoter in T2DM patients compared to controls, particularly at CpG-471—a site crucial for TCF12 binding—highlighting its potential role in T2DM pathogenesis.

## 2. Materials and methods

### 2.1. Sample collection

After receiving approval from the Institutional Ethics Committee, blood samples were collected from 30 newly diagnosed patients with T2DM at the Department of Endocrinology, Second Affiliated Hospital of Soochow University, and 30 healthy volunteers between November 2022 and November 2023. Samples were drawn from the patients’ arm veins using disposable sterile syringes and stored in EDTA-containing tubes to prevent coagulation. Collection occurred after an overnight fast, adhering to the WHO 1999 diagnostic criteria. Patients were informed and voluntarily participated in the study as advised by their physicians.

### 2.2. DNA extraction

DNA extraction was performed using the TIANamp Genomic DNA kit (DP304; TIANGEN, Germany) according to the manufacturer’s standard procedure. Genomic DNA was isolated from 200 µl of the collected peripheral blood samples. The purity of the DNA samples was assessed by calculating the absorbance ratio at 260 nm to 280 nm using an Invitrogen Nanodrop spectrophotometer. The isolated DNA samples were stored at −80°C for further analysis.

### 2.3. McrBC-PCR analysis

McrBC is a GTP-dependent specific nucleic acid endonuclease that acts on methylcytosine-containing DNA and does not act on unmethylated DNA[33]. The DNA methylation level of the region upstream of the KCNJ11 transcription start site was detected by McrBC-PCR. McrBC digestion was performed with at least 200 ng of genomic DNA using the McrBC kit (M0272S; NEB) according to the manufacturer’s instructions. The same amount of DNA without McrBC digestion was used as a negative control. Primers were designed using Primer3, sequences of primers used for McrBC PCR are listed in Supplementary Table 1, 50ul of the PCR amplification system was used and the products were analyzed by electrophoresis on a 1.5% agarose gel.

### 2.4. Bisulfite sequencing PCR (BSP)

DNA Bisulfite Conversion Kit (E3318S; NEB) was applied to DNA bisulfite modification according to the instructions. The total PCR system was 50μL, to which 6μL of bisulfate-modified DNA template, 25μL of 2×PCR buffer, 2μL each of upstream and downstream primers were added, and the final volume was adjusted to 50μL with double-distilled water. The PCR conditions and setting reaction procedures were as follows: pre-denaturation at 94℃ for 3 min, denaturation at 94℃ for 30s, annealing at 55℃ for 30s, and extension at 72℃ for 16s for 35 cycles, and the reaction ended with a final extension at 72℃ for 7 min. The PCR products were electrophoresed in a 2% agarose gel, after which the target fragments were recovered. The pMD18-T vector (6011, TaKaRa) was used for TA clonal ligation, followed by transformation of DH5α receptor cells. Positive clones were selected and sent to AZENTA for sequencing, and the methylation rate was calculated as the number of methylated cytosine (mC) clones/10×100%. The sequencing results were analyzed online according to Web-based Kismeth software (http://katahdin.Mssm.edu/6ismet/revpage.pl)[34].

### 2.5. Chromatin immunoprecipitation (ChIP) assay

ChIP assay was used to validate the binding of the transcription factor TCF12 to the promoter of the KCNJ11 gene. 293T cells were fixed with formaldehyde, and glycine was added for cross-linking. The samples were then placed on ice and sonicated to isolate DNA. Cell lysates were incubated overnight with antibodies (anti-TCF12 antibody(sc-28364,Santa Cruz Biotechnology) or control IgG) at a dilution of 1:50 and then with protein G-agarose beads (#3956088, Merck KgaA, Germany) at 4°C overnight. Mouse control IgG (#3452481, Merck KgaA, Germany) was used as a negative control for the reaction. The bound DNA-protein mixture was eluted and the cross-linking was reversed after several washes. The DNA fragments were then purified and subjected to PCR with the KCNJ11 primer. The PCR products (172 bp) were separated on a 2% agarose gel and visualized on a UV transilluminator.

### 2.6. Bioinformatic analysis

Short reads downloaded from the SRA database with accession SRP274496 were used to produce quality control reports through FastQC (https://www.bioinformatics.babraham.ac.uk/projects/fastqc/) and subjected to low-quality filtering and adapter trimming using EtoKi (https://pubmed.ncbi.nlm.nih.gov/31809257/). Clean data were aligned to the reference genome using Hisat2 (https://pubmed.ncbi.nlm.nih.gov/31375807/), and the mapped results were sorted using Samtools (https://pubmed.ncbi.nlm.nih.gov/19505943/). Transcriptome assembly and counting were carried out using StringTie (https://pubmed.ncbi.nlm.nih.gov/25690850/). Bioinformatic analysis was performed using the R command line, and differential expression analysis was conducted using edgeR (https://pubmed.ncbi.nlm.nih.gov/19910308/). R packages pheatmap (https://cran.r-project.org/web/packages/pheatmap/index.html) were used for visualization.

### 2.7. Quantitative real time polymerase chain reaction assay

Peripheral blood leukocyte RNA was extracted with TRIzol ^TM^ Reagent (15596026CN, Thermo Fisher Scientific), and reverse transcription was performed according to the instructions of the HiScript II Q RT SuperMix for qPCR(+gDNA wiper)(R223,Vazyme) kit. The total volume of the QPCR amplification mixture was 10μL, and 1μL of cDNA template, 5μL of 2*ChamQ SYBR qPCR Master Mix (Q341,Vazyme,), and 0.2μL of upstream and downstream primers were added. The volume was adjusted to a final volume of 10μL with double-distilled water. The PCR conditions and the setup of the reaction program were as follows: pre-denaturation at 95 ℃ for 180s, denaturation at 95℃ for 15s, and annealing and extension at 60℃ for 60s, and the reaction was run for 39 cycles.

### 2.8. Statistical analysis

The data are presented as the mean ± SD. Differences in mean values between two groups were analyzed using the two-tailed t-test. Statistical analyses were performed using the GraphPad Prism version 6.0 (GraphPad Software, USA). *p < 0.05, **p < 0.01, ***p < 0.001 or ****p < 0.0001 were considered statistically significant.

## 3. Results

### 3.1. Clinical characteristics of the study population

To maintain a consistent genetic background and characterize the course of the disease, several potential confounders were considered in this study based on the literature and clinical judgment. Covariates included age, sex, body mass index, smoking status, alcohol consumption status, family history, physical activity, creatinine, total cholesterol, triglyceride, and low-density lipoprotein levels of the included subjects. The data included in this study were categorized into two types of continuous and categorical variables. Continuous variables are expressed as the mean ± standard deviation and were subjected to t-test (normal distribution) or Kruskal-Wallis rank sum test (non-normal distribution) depending on the normality of the distribution; categorical variables were expressed as percentages and were subjected to the χ2 test (**Table 1**). The subjects were divided into a healthy control group and a first-onset type 2 diabetes group according to the WHO 1999 diabetes diagnostic criteria. After the application of strict inclusion criteria, Table 1 shows that the differences between the two groups were statistically significant only for fasting blood glucose, two-hour postprandial blood glucose, and glycosylated hemoglobin(P<0.001).

**TABLE 1.**
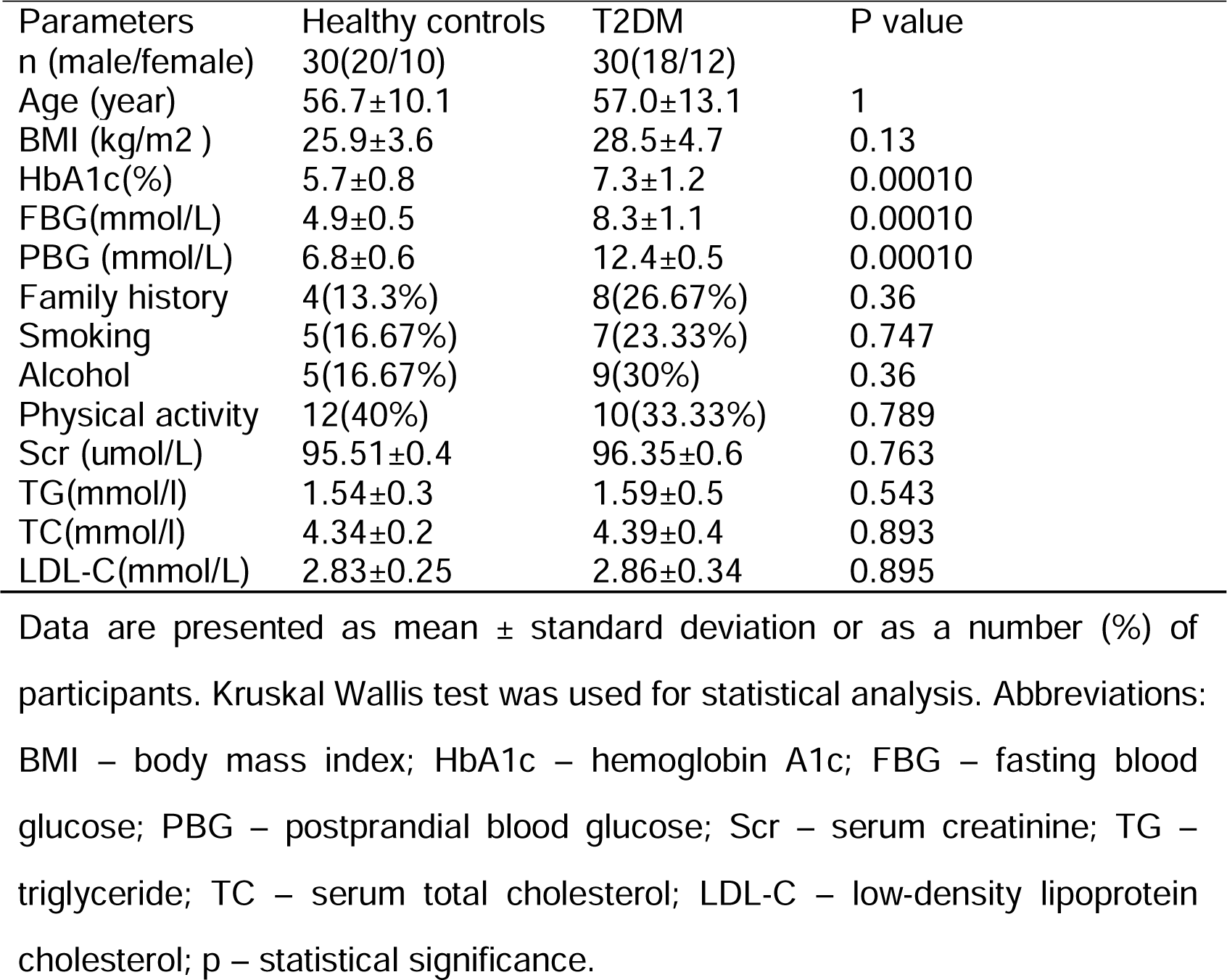
Clinical Characteristics of the subjects.

### 3.2. McrBC-PCR analysis

Agarose gel electrophoresis of the PCR products of diabetic and healthy control samples with the same amount of DNA before and after 1h of McrBC digestion and analysis of gray values are shown in (**Fig. 1**). The results demonstrated that DNA methylation modification exists in the promoter region of the KCNJ11 gene in patients with first-onset T2DM and healthy controls, and that the degree of methylation modification in healthy control samples is likely to be greater than that in patients with first-onset T2DM.

**FIG. 1.**
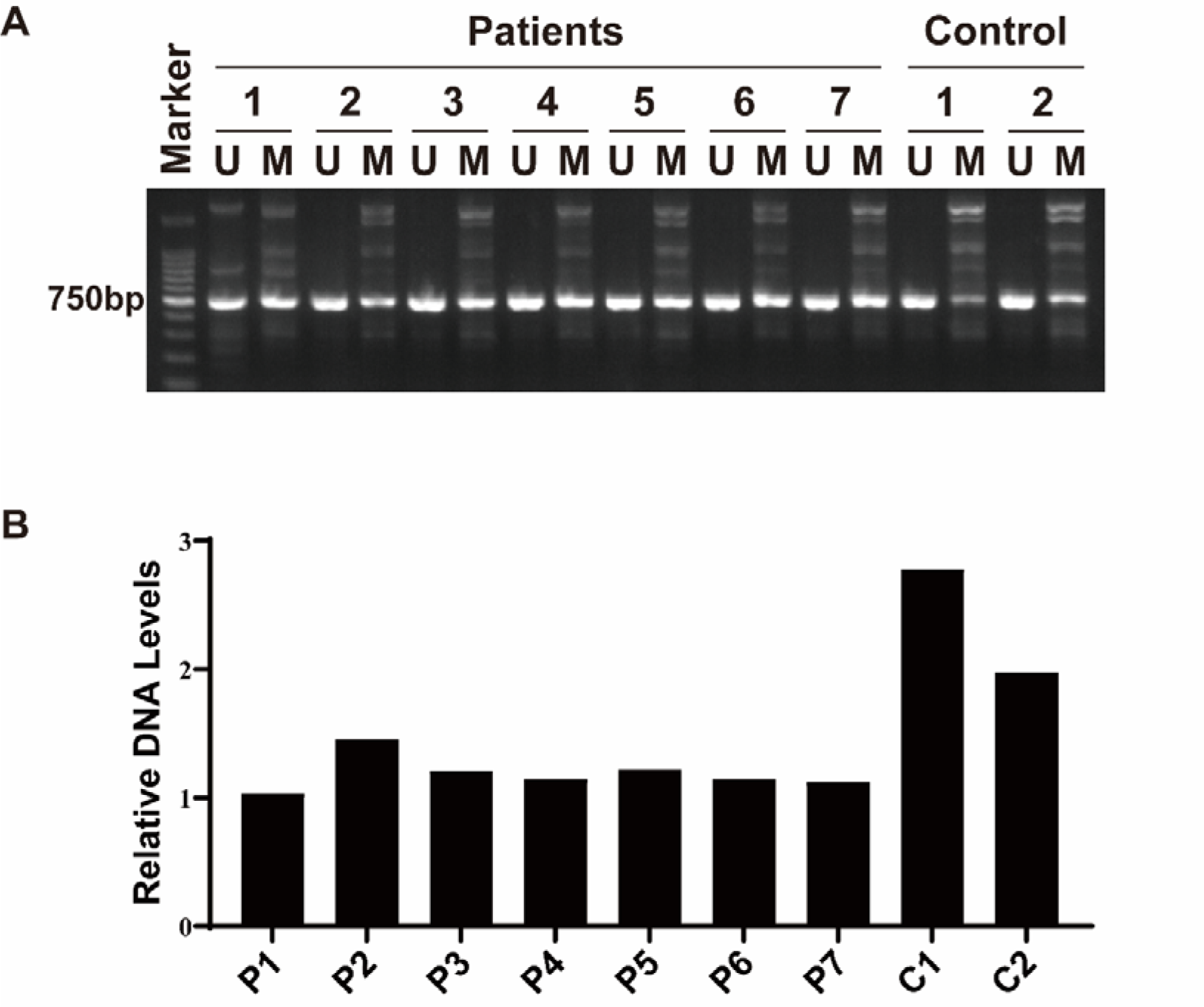
Gel electrophoresis of different diabetic samples and healthy subjects after 1h digestion by McrBC. The same amount of DNA from the same sample on the left side is the uncut control, and on the right side is the digestion for 1h (A); The below panel shows the bar graph of gray value analysis (B). Abbreviations: U – undigested; M – McrBC digested; P – patient; C – healthy control.

### 3.3. BSP results confirm differences in site-specific methylation of the KCNJ11 promoter region between healthy controls and patients with first-onset T2DM

We analyzed DNA methylation at selected locations in this region in the healthy and T2DM groups using the bisulfite sequencing method, as shown in Figure 2B, and the results based on Kismeth software analysis showed an overall decrease in cytosine methylation levels from 22.22% to 3.33%.

**FIG. 2.**
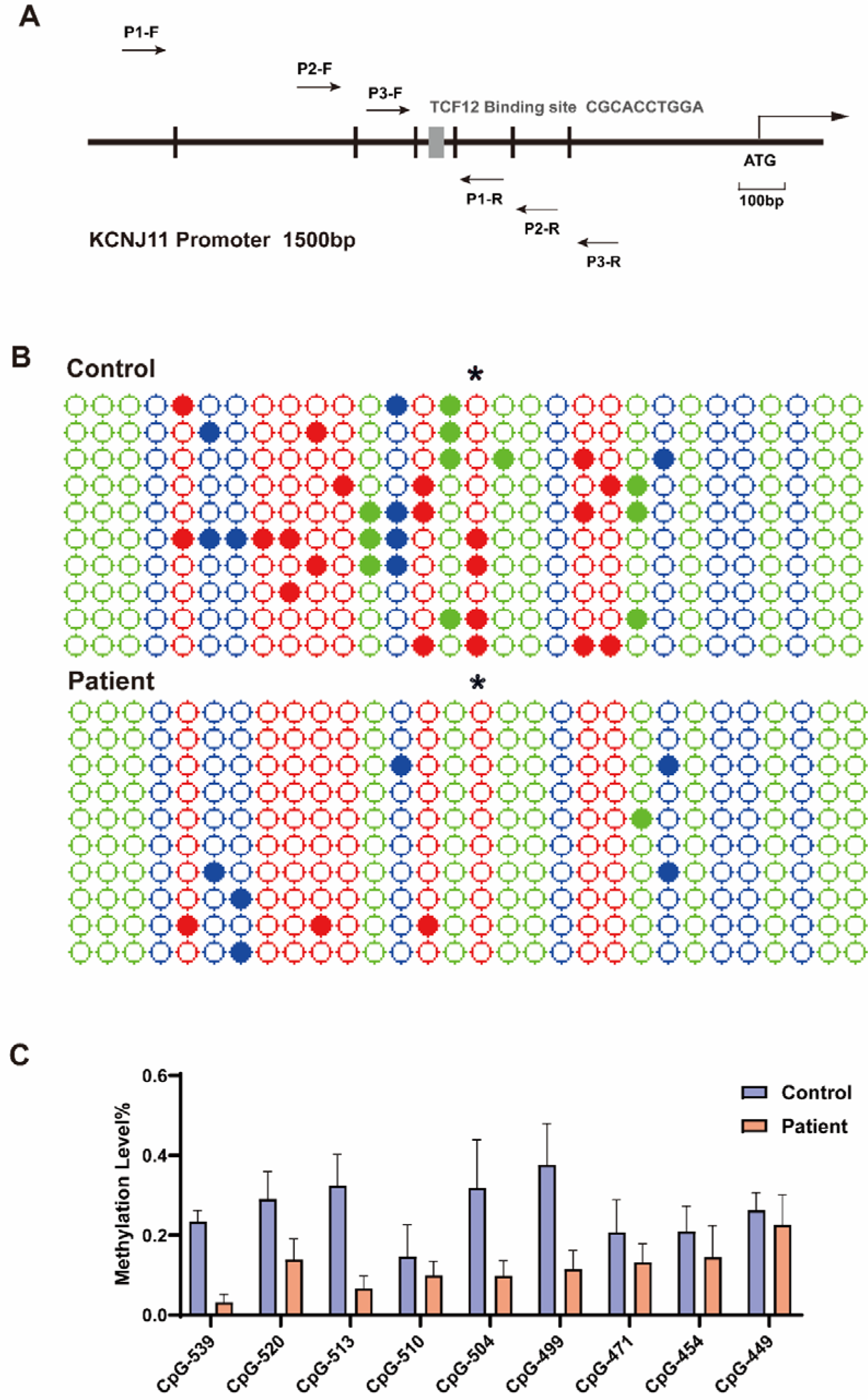
Schematic illustration of the amplification positions of each primer pair in the promoter region of KCNJ11 and the binding site of the TCF12 transcription factor.P1 represents the MCrBC primer (668bp), P2 represents the BSP primer (151bp), and P3 represents the CHIP primer (172bp)(A).Healthy control and T2DM genomic DNA were sequenced for different clones after treatment with sodium bisulfite, and scatters represent cytosines at the positions indicated in the KCNJ11 5’ region, * labeled at the CpG-471 site (B).Histogram of the methylation rate of the KCNJ11 promoter region in the first-episode T2DM group (red bars) and healthy control group(blue bars) (C). Abbreviations: P – primer; F – forward; R – reverse.

Moreover, after bisulfite modification treatment and sequencing of positive clones, it was demonstrated that there were methylation sites at nine sites, including CpG-449, CpG-454, CpG-471, CpG-499, CpG-504, CpG-510, CpG-513, CpG-520, and CpG-539.The methylation ratios of the above loci were significantly lower in patients in the type 2 diabetes group, whereas they were hypermethylated in the healthy normal control group. Moreover, at the CpG-471 locus, the methylation rate was decreased by approximately 7.55% in the T2DM group compared with that in the healthy control group **(Fig. 2C)**.

### 3.4. Binding of differentially methylated sites in the promoter region of the KCNJ11 gene to the transcription factor TCF12

DNA methylation has been extensively studied over the past few decades, and the attachment of methyl groups to the promoter region of a gene might inhibit transcription factor binding, and hence usually reduce transcription; the opposite phenomenon also holds true[35, 36]. The results analyzed by JASPAR and AlphaFold online software showed that the presence of TCF12 transcription factor binds to the promoter region of the KCNJ11 gene near the differentially methylated site CpG-471, which may explain the hypomethylation of the KCNJ11 promoter region in patients with T2DM, which can affect protein expression by enhancing the activity of the transcription factor TCF12 and thus predispose them to developing T2DM **(Fig. 3A)**. CHIP-PCR gel electrophoresis revealed target bands in the input group and anti-TCF12 group but no bands in the IgG or blank group **(Fig. 3B)**, which also confirmed that the transcription factor TCF12 binds to the promoter region of the KCNJ11 gene and may be involved in the methylation changes at specific loci of the KCNJ11 gene, possibly influencing the regulatory pathways involved in pancreatic beta-cell function.

**FIG. 3.**
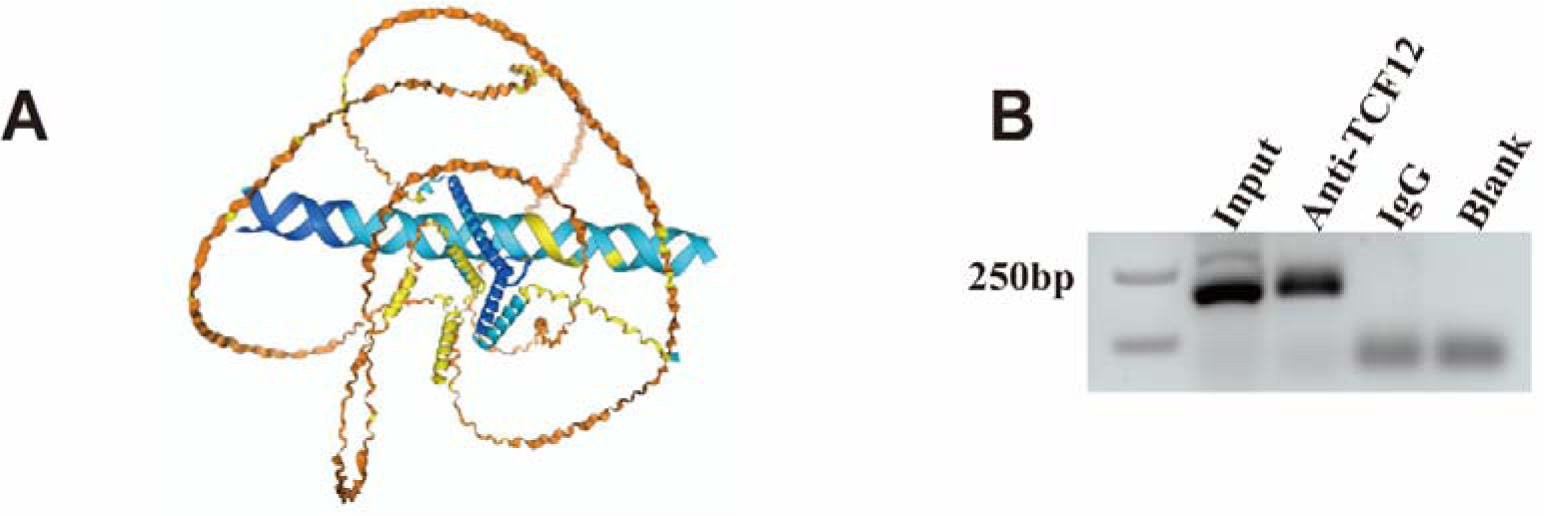
Structural pattern of the binding of the transcription factor TCF12 to the KCNJ11 promoter region demonstrated by AlphaFold software(A).The PCR products of the 293T cells from the ChIP assay were visualized on a UV transilluminator (B).

### 3.5. KCNJ11 expression in human peripheral blood

The RNA-seq data from 12 blood samples obtained from the SRA database, including 6 from healthy individuals and 6 from type 2 diabetes patients, were included in the analysis. Differential gene expression analysis revealed that the KCNJ11 gene was upregulated in the type 2 diabetes group (logFC = 1.478151, p = 0.09869249, FDR = 0.3574878) **(Fig. 4A)**. We then extracted RNA from fresh peripheral blood of enrolled healthy controls and T2DM patients for semi-quantitative analysis of the KCNJ11 gene, and the results demonstrated that KCNJ11 mRNA expression was increased in peripheral blood from patients with T2DM compared with that in peripheral blood from healthy controls (p < 0.001) **(Fig. 4B)**.

**FIG. 4.**
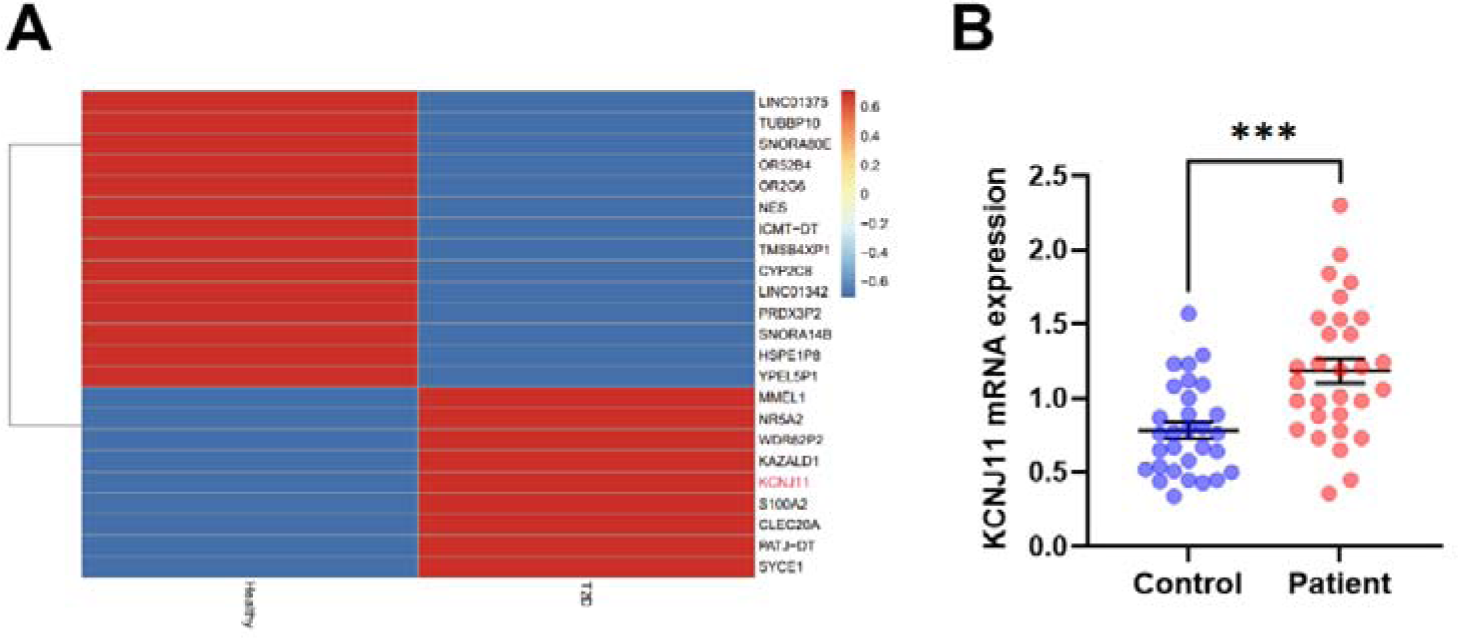
RNA-seq data shows that the KCNJ11 gene is upregulated in the type 2 diabetes group (logFC = 1.478151, p = 0.09869249, FDR = 0.3574878) (A). KCNJ11 mRNA expression levels in human peripheral blood from healthy controls and patients with T2DM. Gene expression was analyzed using quantitative RT-PCR. The results are expressed as the mean ± SD (B).

## 4. Discussion

In this study, BSP-sequencing was utilized to examine methylation levels at specific sites within the KCNJ11 gene promoter in blood samples from individuals at the early stages of Type 2 Diabetes Mellitus (T2DM). Employing bisulfite conversion, a well-established technique for DNA cytosine methylation analysis[37], our results demonstrated distinct differences in methylation patterns at several critical sites between newly diagnosed T2DM patients and healthy controls.

The current literature establishes a clear link between DNA methylation at gene promoters, CpG islands, and proximal gene body sites with changes in transcriptional activity and gene expression[38, 39]. Our findings reinforce this paradigm, revealing significant hypomethylation at pivotal sites within the KCNJ11 promoter in T2DM subjects. This epigenetic alteration likely facilitates increased binding of the transcription factor TCF12, potentially augmenting protein expression and, consequently, T2DM susceptibility. This observation underscores the therapeutic potential of targeting TCF12, not only enhancing our understanding of its regulatory mechanisms but also promoting it as a viable target for T2DM treatment.

Research within the diabetes field is predominantly focused on two approaches: comprehensive scans for differential DNA methylation across the genome[40] and targeted investigations into known diabetes-associated genes using specific primers to pinpoint differential methylation[15, 16, 41, 42]. Our study’s focus on the KCNJ11 promoter methylation contributes significantly to the broader epigenetic narrative of T2DM pathogenesis, paving the way for novel insights into disease mechanisms and therapeutic innovation[43]. The role of methylation in other gene loci related to diabetes progression remains an open question warranting further exploration.

With diabetes prevalence increasing globally, the imperative for innovative therapeutic approaches is more pressing than ever. Future treatment modalities might include epigenetic editing of specific genes[44], a strategy supported by emerging research advocating for targeting the enzymes that modify epigenetic landscapes in tissues affected by diabetes[45, 46]. Moreover, epigenetic changes in diabetic patients have been increasingly associated with severe vascular complications, such as retinopathy, stroke, and myocardial infarction[47–49]. Identifying epigenetic biomarkers, readily detectable in blood and derivative from target tissues, could profoundly impact clinical practices by facilitating easier analysis in diabetic and at-risk populations. However, the discovery of epigenetic markers with superior predictive strength over existing diabetes indicators remains a critical research need.

In summary, our study has identified significant epigenetic modifications in the promoter of the KCNJ11 gene in patients with T2DM, highlighting a robust link between epigenetics and the pathophysiology of diabetes in humans. These epigenetic markers offer promising avenues as biomarkers for predicting T2DM, gauging the risk of vascular complications, and assessing the efficacy of therapeutic and lifestyle interventions, thus heralding a new era of precision medicine in diabetes care.

## Data Availability

All data produced in the present work are contained in the manuscript

## Acknowledgements

This research was supported by grants from the National Natural Science Foundation of China (82371544,82070814); Suzhou Healthcare Science and Technology Innovation Program (SKYD2022030); Open project of Jiangsu Health Development Research Center (JSHD2022035); A Project Funded by the Priority Academic Program Development of Jiangsu Higher Education Institutions.

